# The development of a World Health Organization transdiagnostic chatbot intervention for distressed adolescents and young adults

**DOI:** 10.1101/2025.02.19.25322432

**Authors:** Dharani Keyan, Jennifer Hall, Stewart Jordan, Sarah Watts, Teresa Au, Katie S. Dawson, Rajiah Abu Sway, Joy Crawford, Katherine Sorsdahl, Nagendra P Luitel, Anne M. de Graaff, Heba Ghalayini, Rand Habashneh, Hafsa EL-Dardery, Sarah Fanatseh, Aiysha Malik, Chiara Servili, Muhannad Faroun, Adnan Abualhaija, Ibrahim Said Aqel, Syed Usman Hamdani, Latefa Dardas, Aemal Akhtar, Richard A. Bryant, Kenneth Carswell

**Author notes:** Correspondence to: Dharani Keyan, PhD School of Psychology, University of New South Wales, Sydney, NSW 2052 Australia.

## Abstract

**Background:** Common mental disorders are prevalent in young people in low- and middle-income countries (LMICs). Digitally delivered interventions have the potential to overcome many structural and psychosocial barriers to mental health care. Chatbots have been proposed as one potentially acceptable and feasible method that may increase engagement. Yet, there is currently limited evidence for their efficacy in reducing psychological distress. This paper summarises the development of a World Health Organization digital psychological intervention for young people experiencing impairing psychological distress, developed in line with Human Centred Design (HCD) principles.

**Objective:** This study refined and adapted a chatbot intervention initially developed for adolescents aged 15-18 years that was completed in consultation with end-users in this age group (N =236), community members (N =73), and psychology intervention experts (N =9) across varied settings. The purpose was to create an adaptation fit for use by young adults aged 18-21 years experiencing psychological distress in Jordan.

**Methods:** The current study followed a limited user-centred design process involving focus groups and key informant interviews with stakeholders including young adults aged 18-21 years (N =33), community members (N= 13), and psychology intervention experts (N= 11). Iterative design development occurred throughout the cultural adaptation and refinement process.

**Results:** There was a clear preference for a chatbot based intervention that included interactions with fictional characters with relatable problems. The chatbot content followed a transdiagnostic model that addressed common problems including low mood, stress and anger with reference to vocational, familial and interpersonal stressors that the target population commonly faced. It followed a non-AI decision tree format with multiple sessions and was designed to be adaptable for use in different countries with different populations and software systems. Prototype versions of the chatbot were well-received by adolescents (15– 18-year-old) and young adults (18–21-year-old).

**Conclusions:** This is the first report of the development of a chatbot intervention for adolescents and young adults in LMICs that was designed using a HCD framework. Systematic end-user engagement through all phases of the research aimed to make this intervention acceptable and useable for adolescents and young adults in a wide variety of settings. The chatbot is currently being tested in randomised controlled trials in Jordan and Lithuania.

## 1 Introduction

Common mental disorders including anxiety and depression are among the leading causes of disability globally amongst young people aged 10-24 years (Solmi et al., 2022; Vos, 2020). Exposure to poverty, potentially traumatic events, and other adverse circumstances can negatively affect the mental health of young people, particularly in low- and middle-income countries (LMICs) (Kieling et al., 2011). Approximately 14% of young people (aged 10-19 years) around the world live with a mental disorder (World Health Organization, 2022). In 2019, depressive- and anxiety disorders were ranked as the 4^th^ and 7^th^ causes of disability in young people aged 10-24 years globally (GBD 2019 Diseases and Injuries Collaborators, 2020). Many young people report fears of social exclusion, stigma and discrimination that deter them from seeking help and accessing mental health services (Renwick et al., 2022). Psychological interventions for young people in LMICs, including children, adolescents and young adults show some, but limited effectiveness for reducing symptoms of common mental health conditions (Barbui et al., 2020; Purgato et al., 2018).

One reason for this may be that many of these programs have been heavily conceptualised within a ‘top-down’ framework. The active participation of young people in the conceptualisation and design of psychological interventions in LMICs may help to make them more effective, accessible, and appealing.

Digital mental health interventions may be one way of providing evidence-based support, especially in places where resources and access are limited. Specifically, digital approaches to mental health care may be perceived as less stigmatising as they offer anonymity and may reduce social barriers to seeking support. Online applications show promise for treating common mental health problems (Firth et al., 2017); yet, many suffer from low completion rates and early disengagement. An independent participant data meta-analysis of self-guided online interventions for depression found that less than one quarter of adults complete all the treatment content (Karyotaki et al., 2015). The integration of human support (guided self-help) may help to mitigate this adherence problem (Baumeister et al., 2014). For example, the provision of support from a helper on the use of interventions tends to yield somewhat increased engagement (Gan et al., 2021). In turn, this increased engagement may be associated with improvements in mental health (Gan et al., 2021).

Conversational agents or chatbots provide another type of digital delivery that can provide information in a potentially more engaging manner. Chatbot platforms in youth mental health have typically utilised either a pre-programmed approach using decision tree logic (Ly et al., 2017), or machine leaning methods (Fitzpatrick et al., 2017) to simulate a messenger style conversation wherein a user receives instruction on therapeutic skills. There is some evidence to suggest these are acceptable and feasible for youth experiencing common mental health conditions such as depression and anxiety (Daley et al., 2020; Fitzpatrick et al., 2017), there is less evidence to support their longer-term effectiveness in reducing distress (Vaidyam et al., 2019).

The scope to automate much of the content in chatbot platforms may provide a novel opportunity for improving availability of evidence-based mental health care in resource constrained LMICs that are affected by both access and contextual barriers, especially with the increasingly prevalent use of mobile devices across many LMICs (Naslund et al., 2017). With an increasing proportion of populations in LMICs connected through mobile technologies, and with more than 80% of all people with common mental health conditions living in LMICs (World Health Organization, 2022), there is an opportunity to reach vulnerable individuals across wide-ranging settings using digital interventions. Part of the World Health Organization’s (WHO) initiative on increasing access to potentially scalable psychological interventions has involved developing transdiagnostic interventions that use digital technology. One of these, the WHO *Sustainable Technology for Adolescents and youth to Reduce Stress* (STARS) intervention is a chatbot based intervention for youth experiencing psychological distress. This paper provides a detailed outline of the extensive development process undertaken in the initial design (Hall et al., 2022) and subsequent revision, and adaptation, both of which comprised a human centred design (HCD) approach involving iterative consultations with end-users.

## 2 Background

### 2.1 Overall approach to design

To address many of the problems identified in prior research on digital mental health interventions for adolescents and youth, one of the main goals of the project that led to the development of STARS was to apply human centred design (HCD) principles. This meant there was no fixed idea of whether the intervention would be a smartphone app, website, chatbot or something else. Instead, the approach combined a top-down process (e.g., drawing on available evidence of effective psychological interventions and techniques) with an extensive bottom-up process that actively involved young people (including both adolescents aged 15-18 years, and young adults aged 18-21 years) and other key stakeholders across the phases of designing the digital intervention. Essentially, this approach aimed to find ways that the evidence-based techniques could be delivered in a highly engaging way. In part, HCD consists of iterative stages of development to ensure that the end-product is strongly grounded in the context of the users who ultimately will use it (Scholten & Granic, 2019; Taylor Salisbury et al., 2021). To do this, HCD employs a user-centred approach that includes testing design assumptions or questions with multiple sources of information (i.e., empirical research, feedback from experts and end users, prototype testing) to understand the person within their context, which is hypothesised to make a given intervention more meaningful and in turn more useful and engaging to end users. HCD approaches have been used previously to support the implementation of health interventions in low-resourced settings (Lyon & Koerner, 2016). To a degree, HCD is an extension of commonly applied user-centred iterative processes that guide the cultural adaptation of interventions (Abi Ramia et al., 2018; World Health Organization, 2024). STARS was developed across two distinct development and testing stages that are detailed below (see Figure 1).

**Figure 1.**
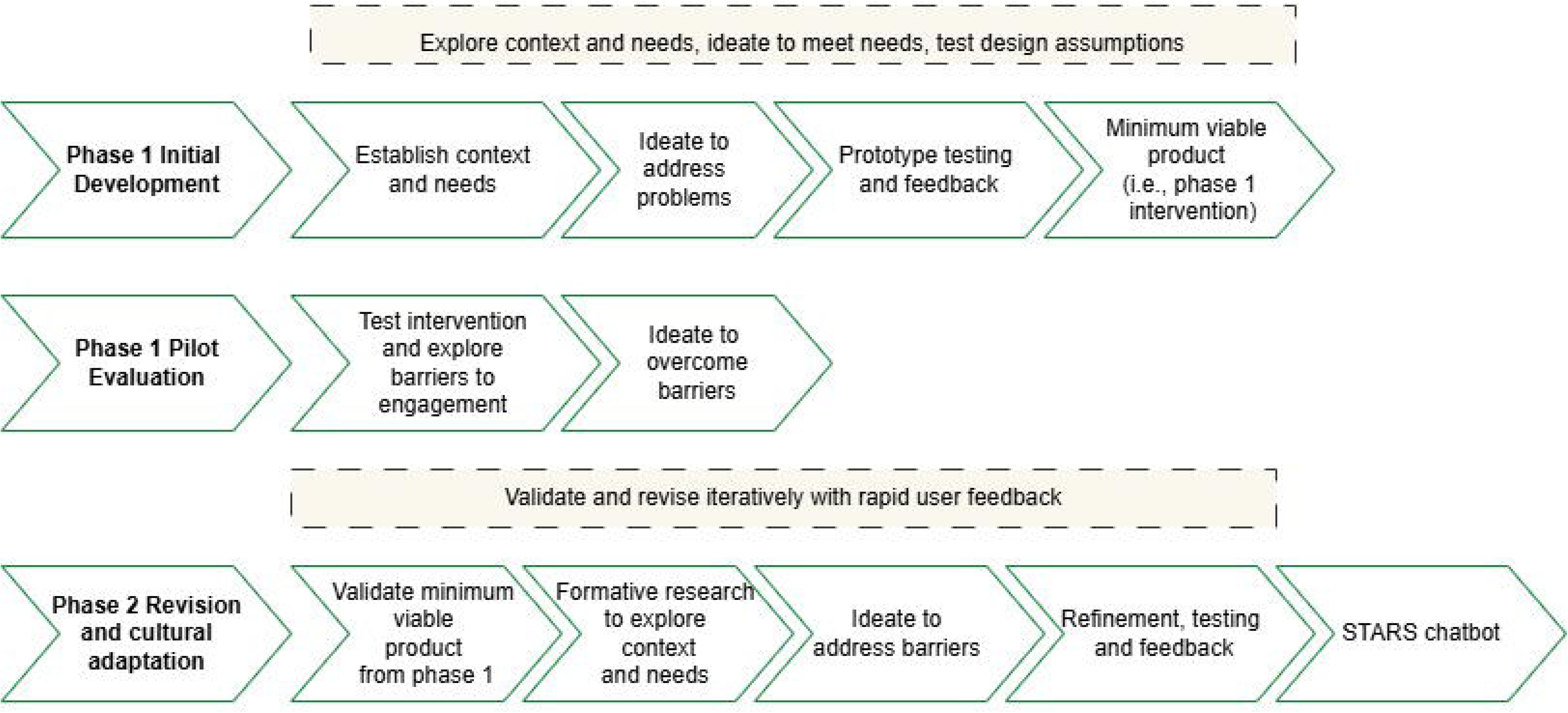
The HCD framework underpinning the development of the STARS chatbot.

### 2.2 Previous work: Phase 1 development

The initial target population was young people aged 15-18 years of all genders, reporting significant psychological distress, living in urban areas and from a low-to-middle income socioeconomic status. Young people in this age range from varied settings took part in the intervention development activities including 59 from Pakistan 66 from the West Bank and Gaza, 36 from Jamaica, 42 from South Africa and 33 from Nepal. Additionally, individuals with regular contact with an adolescent were also included in some activities including 23 from Pakistan, 13 from Gaza and West Bank, six from Jamaica, 22 from South Africa, and 11 from Nepal. They consisted of parents and caregivers, teachers, counsellors, and health professionals trained on mental health. Interviews with adolescent and community members across all sites took place in contexts of substantial adversity which included ongoing conflict, crime, gang violence and poverty. Different settings reported different causes and levels of adversity. Interviews were conducted in a range of native languages including Afrikaans, Arabic, English, isiXhosa, Nepali, Patois, and Urdu.

Activities were conceptualised in iterative cycles of stages following HCD principles. Stage one focused on (1) establishing the context surrounding young people and their needs including their daily lives and hobbies, perceptions of distress, help-seeking behaviour, technology use, current ways of coping, and motivation to engage in interventions. The second stage involved (2) developing ideas to overcome identified problems. This included co-creation activities where fictional personas developed from the previous stage were reviewed to help generate ideas to inform development of the intervention. The final stage consisted of (3) building and testing prototypes where participants were presented with multiple basic prototypes (i.e., content from a module, and methods to deliver this content), each demonstrating an alternative method of achieving the objective that was being tested. Participants provided feedback which informed the next update of the prototypes. This final phase was iterative in that multiple of rounds of prototype testing were conducted until a final product was developed.

The range of activities were conducted in formats including interviews and focus groups. All participants provided informed consent, and, in some settings, non-cash incentives were provided (e.g., drinks, food, pens, supermarket- or phone vouchers).

Facilitators had prior experiences in working with adolescents, were from a similar cultural background. The WHO-led design team comprised of an HCD expert (SJ) that supported the initial development alongside other authors with a background in clinical psychology (JH, TA, SW, KD, KC). Interviews with key stakeholders including end users (i.e., adolescents), community members, and psychological experts were conducted to understand the perspectives and context of the target group. Finally, literature reviews and consultations with global experts (n = 9) were conducted by WHO. These consultations focussed on expert opinions about online interventions for adolescents, ways to adapt interventions for this target group, opinions relating to what works with this population and recommendations for the proposed intervention.

### 2.3 Previous work: Phase 1 Synthesis

Key themes were identified on topics including the use and availability of technology, emotional health, needs, motivations barriers and enablers for adolescents. These were then used by the design team to develop fictional personas, that were reviewed by partners in the different sites and in design workshops where adolescents and community members provided feedback on how well the fictional personas represented adolescents in their community. The outcomes from this process were used to guide the development of multiple prototypes that were updated and tested across iterative cycles.

#### 2.3.1 Establishing the user context and presenting needs

Phase 1 activities showed that adolescents experienced difficulties with stress and low mood related to challenges including school and/or university, and in familial and romantic relationships. Internalising problems (e.g., withdrawing from close others, loss of concentration, feeling lonely), externalising problems (e.g., anger outbursts, fighting with others) and somatic problems (e.g., tightness or tension, headaches) were seen as important to address. Stigma was an often-mentioned barrier to help seeking, with experts emphasising the need to normalise experiences of distress to decrease feelings of shame about help-seeking.

Adolescents in the communities were familiar with using a smartphone, ownership varied, but most could access through a shared family phone. Memory space on phones and access to substantial internet data was often limited and a barrier to downloading apps. Devices were used to learn, watch videos, and play games.

#### 2.3.2 Ideas to overcome problems and address needs

Activities highlighted an interest in a “blended” approach involving the use of videos, stories, games, individualised content that catered to interests (e.g., football, dancing), interactive stories with ‘real-life’ characters (i.e., fictional personas), and a sense of control over the direction of the overall narrative (e.g., “choose your own character story”).

Reportedly helpful features of commonly used digital applications included immediate rewards, control over the flow of a story or game, social networking, avatars and use of conversations.

This information was used to develop fictional personas. Information on each persona included age (e.g., 16-year-old), occupation (e.g., student), family context (e.g., lives at home and shares a room with mother and 2 young sisters), technology use (e.g., has an android phone that is shared with sisters; uses it to listen to music, watch videos) internet access (e.g., mother buys data, and if not, access Wifi at home and school), perceptions of wellbeing (e.g., often feeling sad or stressed), needs (e.g., watches videos due to poor concentration), motivations (e.g., motivated to concentrate, so can study and get better grades), barriers (e.g., being worried that others will find out about emotional state), and enablers (e.g., wants emotional support) to engagement. Key assumptions about user preferences based on these findings were developed and included “*young people want a psychological intervention delivered by a peer*”, and “*young people prefer the intervention to be in a conversational format*”. These were then tested through further activities such as prototype testing.

Literature reviews and expert interviews suggested the use of a transdiagnostic approach. The benefit of this approach is that core factors underlying mixed emotional and behavioural difficulties in adolescent and youth populations can be targeted (Weisz et al., 2015). There is emerging evidence for these approaches with youth with common mental health problems (Jeppesen et al., 2021; Michelson et al., 2020) which supports the existing evidence base for transdiagnostic interventions in LMICs (Murray et al., 2018; Murray & Jordans, 2016).

#### 2.3.3 Prototype testing of content and user feedback

Prototypes of potential transdiagnostic psychological content (e.g., psychoeducation, relaxation exercises, goal setting exercises) delivered in different ways (e.g., presented using chatbots, videos, audios, written information sheets presented in an app) were developed and tested with adolescents across the different sites (N=121). Broadly these prototype tests showed a preference for a diversity of different psychological techniques (e.g. some adolescents preferred breath-based activities for stress management whilst others preferred grounding techniques); the relevance of activities such as goal setting and understanding what to do to feel better; the need for relatable and diverse delivery techniques to address preferences (e.g., use of pictures, videos, relatable techniques) and different situations (e.g., interpersonal challenges, vocational difficulties); and the need to individualise and provide some choice in the intervention. Crucially, this feedback was supported by the views of experts.

There was a clear preference for chatbots which were seen as the most engaging approach and a resounding preference for the use of videos to present tools and techniques, with extensive written information being broadly disliked. The most popular format was a session length between 5 and 10 minutes with information presented in a story format (e.g., a fictional story of someone using deep breathing to relax before a school exam). The ability to choose from multiple fictional stories of characters with relatable problems and the ability to actively engage with these stories, such as suggesting ‘what a character should do next’ were popular.

### 2.4 Phase 1: Minimum viable product (MVP)

This process of iterative prototype testing and review led to the development of the first version of STARS (Hall et al., 2022). This chatbot addressed key issues such as stress, anxiety, sadness, anger and interpersonal problems. This was conceptualised within a transdiagnostic framework and did not use diagnostic terms in order to reduce stigma. Figure 2 represents the transdiagnostic pathway of the MVP. Users follow the same pathway through the chatbot, but content is individualised to an extent with choice over which psychological technique to learn (e.g., deep breathing or grounding), and which stories to follow (i.e., fictional personas with different distress presentations). It was written in an engaging and friendly format.

**Figure 2.**
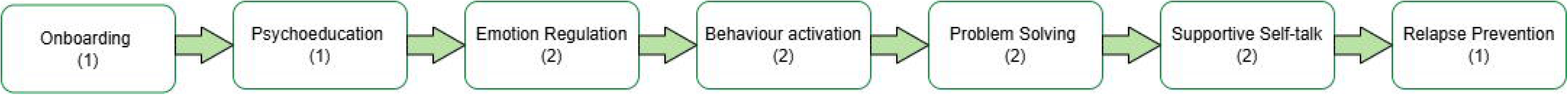
Phase 1 – Content of STARS. Numbers in parentheses refer to number of sessions comprising each module.

### 2.5 Phase 1: MVP evaluation

Twenty-six adolescents aged 15–18-years in South Africa tested the MVP iteratively over three cycles with updates made after each round (unpublished). Extensive feedback was provided on all sessions of the chatbot. Overall, the content was well received. Respondents validated some key aspects of the chatbot including the chatbot “voice” of a trusted older friend (named “Chommie’) was perceived to be friendly, relatable and readily understand the experiences of teenagers. The interactive fictional persona stories were relevant and relatable to adolescents, yet required some changes to make them more specific and realistic; and that video usage (e.g., use of animated GIF to assist in a deep breathing exercise, an animated explainer for a problem management strategy) was helpful. Feedback suggested that the use of pre-defined reply options instead of free text options was experienced as restrictive and interrupted the flow of conversation. Participants wanted opportunities to freely express replies in addition to pre-set reply options.

Improvements suggested including more references throughout the intervention to goals set in earlier sessions; making psychoeducation content less theoretical and easier to read; more support with the behavioural activation activity of breaking down a larger activity into smaller steps; and more needed to be done to support users with practising each of the skills in their own lives. Giving the end-user choices was frequently requested and this has been linked to increased user engagement with digital mental health interventions (Borghouts et al., 2021; Fleming et al., 2017). Examples of providing choices from existing literature include goal setting and selection of psychological techniques (Yardley et al., 2015). Adherence rates also appear to be improved by: the use of non-judgmental language that aligns with the user’s social context; human guidance; and the ability for the user to interact and provide inputs (Kelders et al., 2012).

The design team considered these findings and developed a range of possible ways to improve the chatbot. Ideas for improving the chatbot included the use of a standardised module structure to support individualisation with increased access to summary content (e.g., quizzes) and choice of revisiting or starting a new session. The use of ‘emotion check ins’ at the beginning of sessions where a user could then choose to either repeat content (e.g., a stress management exercise if feeling distressed) or be directed to a different part of the intervention that suited their needs was also identified as a possible way to increase engagement, as was greater use of character stories with more user interaction. The use of summary sections that reinforced core messages was also identified as a strategy. Finally, incorporating human guidance as part of the chatbot intervention was considered within the context of literature evidencing probable increased effectiveness of guided digital mental health interventions in LMICs (Cuijpers et al., 2022; Karyotaki et al., 2021).

### 2.6 Knowledge gap and rationale

WHO aimed to test the phase 1 chatbot intervention in a cluster randomised controlled trial in schools in Jordan. However, due to the impacts of the COVID-19 pandemic, the initial focus on 15–18-year-olds in schools was adjusted to 18–21-year-old youth in the community in urban environments in Jordan. As such, further development and refinement of the phase 1 intervention for this population was the focus of the current study. This involved similar methodologies to the first round of HCD testing, but the process and extent of testing was more limited.

## 3 Methods

In the current study, young adults (aged 18 – 21 years), community members and psychological intervention experts in Jordan were firstly presented with the phase 1 intervention to validate relevance, acceptability and understanding of content (N=17). They were later provided with prototypes showing new features (N=22), all of which were designed based on phase 2 validation activities (N=18), synthesis of findings from phase 1 piloting, and desk research of self-help interventions available in the literature. This iterative process (i.e., phase 2) was led by the current study research team (DK, RH, SF, KC).

### 3.1 Phase 2: Revision and Cultural Adaptation

#### 3.1.1 Context, presenting needs and barriers to engagement

The phase 1 intervention was translated into Arabic and reviewed by 18–21-year-olds who were subsequently interviewed in a focus group discussion using cognitive interviewing, a technique commonly used in the adaptation of interventions (World Health Organization, 2024). The aim of this was to inform further refinement and cultural adaptation of STARS. Groups of youth (n=7), community members (n=5) and psychological intervention experts (n=5) received the content in Arabic and English in both chatbot- and written script formats. They were asked questions about whether it was relevant, understandable, and acceptable following a semi-structured interview guide. Conversational content was selected to purposefully include components that were hypothesized to be less acceptable, and relevant to the target group of 18–21-year-old youth, such as going swimming in Jordan, or character stories relating to school. Participants were asked to offer alternatives to the less acceptable, and relevant content and were asked how they would make the content more understandable for the target population.

Additionally, two focus groups comprising of 18–21-year-old males (n=8) and females (n=10) were conducted to gather insights on their daily routines, engagement in activities, perceptions of distress, ways they connect with others, experience enjoyment, achieve goals and seek support. Participants were also asked about their technology use, currents ways of coping, and motivation to engage in an online chatbot. Questions also explored possible solutions to barriers to engaging with an online intervention identified in phase 1.

#### 3.1.2 Expert read-through and literature review

The phase 1 intervention scripts were reviewed and updated by a clinical psychologist (DK) on the basis of findings from the phase 1 pilot evaluation in South Africa (unpublished) and phase 2 formative research results (i.e., section 3.1.1). Updated scripts were in turn reviewed by young people (n=8), local community members including service providers at the local implementing organisation (n=8), psychology intervention experts (n = 3), and the phase 2 WHO-led development team (DK, AdG, KC; n=3). This was complemented with (i) a non-structured literature search on digital mental health interventions in LMIC settings; and (ii) internal desk review conducted by the WHO team on challenges with using digital interventions in Jordan. Both aimed to further explore key issues raised in focus group discussions with youth.

## 4 Results

### 4.1 User context and presenting needs in Jordan

Young adults (18–21-year-olds) validated the need for support with common mental health problems including sadness, anxiety, anger, and interpersonal problems which arose in the context of distress related to secondary school exams, engagement with university, and a lack of employment opportunities. Reported hobbies included sport, household chores, arts and crafts, cooking, and self-care. Experiences of stigma towards mental health difficulties were commonly reported, where community members reinforced this issue as a considerable barrier to access of mental health services. Experts reported on the importance of normalising challenging emotions, feelings of stigma and related shame expressed by young adults.

Current ways of coping with distress included externalising (i.e., yelling, throwing things) and internalising (i.e., keeping to oneself, sleeping, ignoring the problem) reactions. Young adults expressed some preference for a learned or wise chatbot character (e.g., friend, family friend). They reported liking specific activities that offered connection with others (e.g., helping friends and family), enjoyment (e.g., picnics, dancing) and sense of achievement (e.g., trying new activities). Most young adults were comfortable with online learning (e.g., phone, chatbots) but expressed some concerns about privacy, fears of hacking, and the need for password protection for mental health content.

### 4.2 Validation of MVP and ideas to address needs of young adults

Young adults, community members and experts reviewed the phase 1 intervention and discussed specific methods, including those reported above, which informed the revision and adaptation of content in phase 2. Overall results validated prior findings from phase 1. First, young adults made suggestions to the framing of the intervention in ways that would increase their acceptability. This included specific feedback on revising tone (e.g., a reduced number of emojis) and language style to be more mature; providing the user with the experience of learning from a trusted source that could guide them in learning key skills and cultivate the idea that their coping behaviours are changeable. Second, young adults recommended further personalisation by referring to goals set earlier in the intervention throughout the intervention. Third, the importance of being given some control over the method of learning was emphasized (e.g., choice over completing a stress management exercise as an audio or written exercise). Fourth, young adults wanted relatable stories that matched their context including user testimonials that may facilitate their learning of new strategies. Fifth, all groups supported the need for the chatbot to build agency and competence in skill use. Suggestions included the need for specific and positive feedback (within pre-set reply options) to users following learning of strategies.

### 4.3 MVP refinement and user feedback

Prototypes of new content and features were developed and presented to respondents for feedback. Participants identified with the pre-set goal setting domains (i.e., learning, relationships, mood, developing self) established in phase 1 intervention and offered suggestions for improvement. When presented with the new ‘mood check in’ feature at the start of sessions, participants requested the chatbot to acknowledge their perseverance with the sessions during times of poor mood. Participants suggested ways to increase their engagement with fictional characters, specifically, to be reminded of the experience of speaking to a peer instead of the automated chatbot (e.g., say *‘you can read stories from other young people’*) when presented with the choice of character stories. There were also specific suggestions to increase engagement with character stories, such as inert quick-reply options (e.g., ‘*unsure*’) to be replaced with more active responses. Participants wished to link physical symptoms to signs of stress (e.g., headaches) and happiness (e.g., energetic) during the session on understanding emotions. Participants reported that being reminded that stress can be beneficial when managed well helped to reduce stigmatising attitudes related to managing emotions. In the ‘practice check’ section of the second behavioural activation session, participants recommended increasing their accountability by having the bot refer to specific activities that they had chosen in the previous session. Participants wished to be encouraged by the bot to practise learned skills and asked for revisions to pre-set quick-reply options. Given the feedback on the need for practice of skills, participants were provided with a new ‘*Helping session’* with the opportunity to guide a fictional character to engage in learned strategies to manage a given problem.

## 5 Phase 2: Current STARS chatbot

The content of the STARS chatbot developed through phase 2 is presented in Table 1. Screenshots of the current content of the chatbot are presented in Figure 3. Together, these show the core intervention as conceptualised from this multi-year field testing and user feedback across phases 1 and 2. The proposed psychological intervention (10 sessions across five weeks) consists of a pre-programmed conversational agent (chatbot, called ‘Salam’ in Jordan) that utilises decision tree-logic through text messages to deliver psychological content. In particular, the content is delivered in a conversational format with users being able to select from a range of text responses to respond to the chatbot. This provides the feeling of choice, yet the content provided to all users is ultimately the same. User preferences are accommodated through some choice over notifications and content delivered to them (e.g., which fictional persona they choose to interact with or which relaxation strategy they choose to try out).

**Figure 3.**
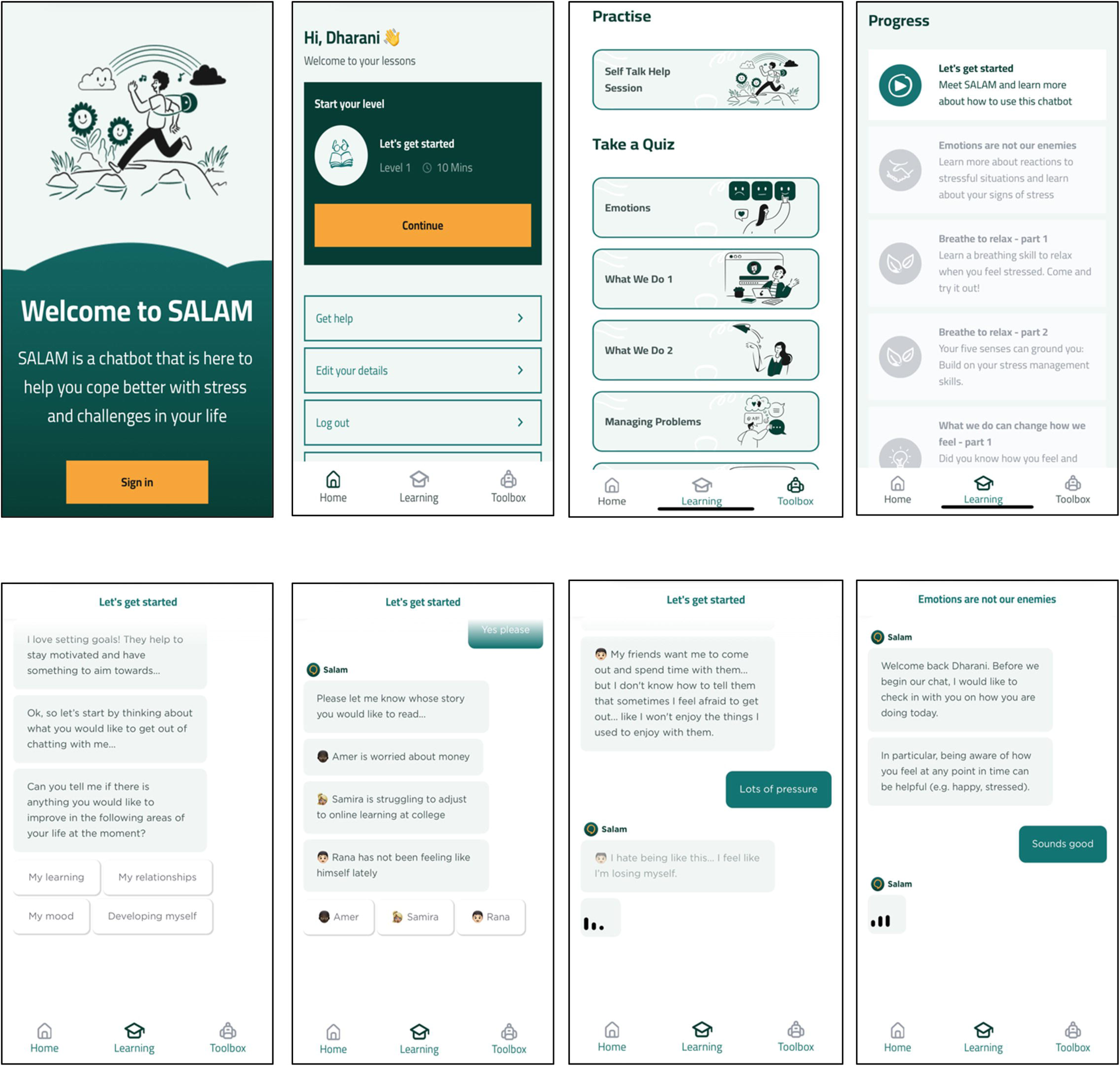
Sample screenshots of the STARS chatbot designed for young adults (aged 18-21 years). The top row (left to right) of wire frames includes screens for the signup page, dashboard, toolbox with practise and quizzes, and progress page. The bottom row of wireframes (left to right) includes goal selection, fictional persona selection, and conversational formats with personas.

**Table 1.**
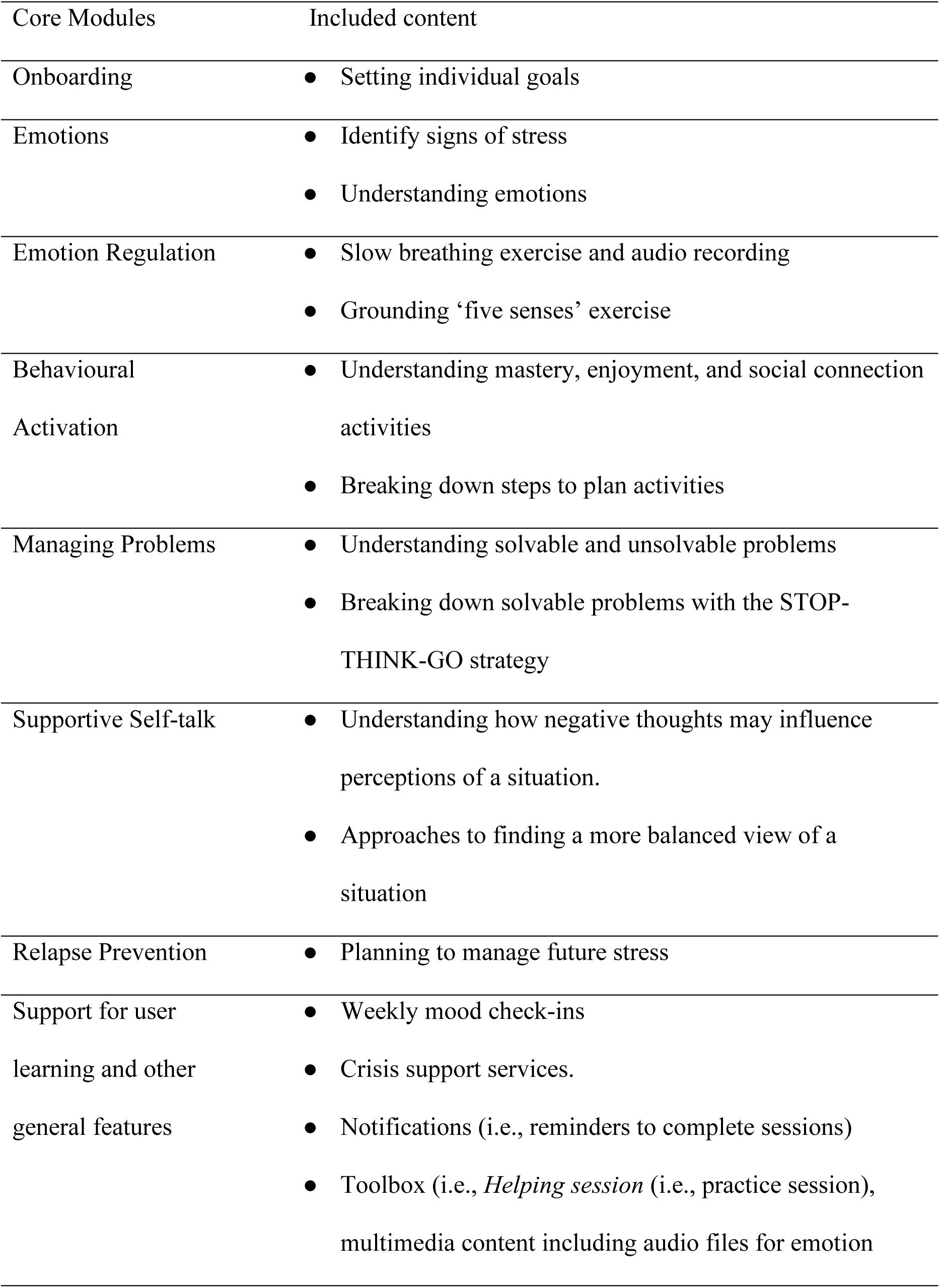
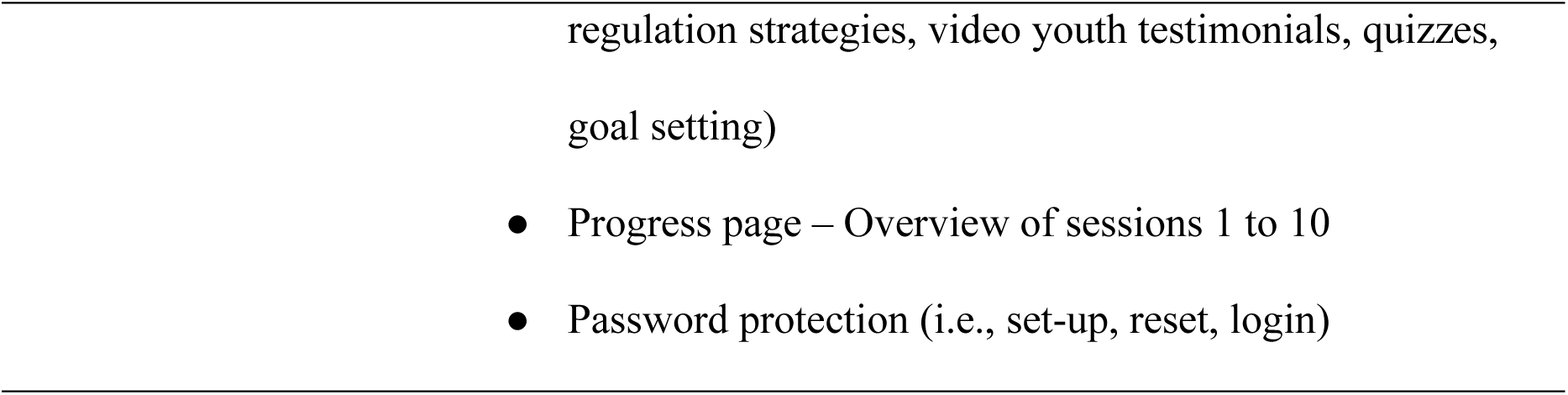
STARS chatbot content.

The chatbot content was made relevant for young adults within an urban environment reporting experiences of stress in relation to university, unemployment, finances, romantic and familial relationships. Fictional personas have been framed to be de-stigmatising of youth experiences of stress and adversity and to normalise different presentations of distress.

Conversations with the chatbot are designed to give the user the experience of feeling like they are speaking to a person (although they are made aware in the initial session that this is a pre-programed chatbot). Chat sessions are longer (i.e., 15-20 mins) than the preferences outlined in Phase 1, and included videos, text, audio clips and activities to cater to different learning styles when communicating core psychological techniques. Additionally, practice sessions, quizzes and re-cap messages throughout interactions with the fictional persona are designed to reinforce learning.

The psychological content follows a transdiagnostic cognitive behavioural therapy (CBT) framework to address the broad mental health needs reported by youth in target sites. At the beginning of each session, users are provided with a ‘mood check-in’ and asked to rate their mood on a 5-point scale (e.g., very bad to very good). When the user gives feedback of feeling less than ‘good’, the bot provides validation prior to continuing with core content.

### 5.1 Core modules of STARS

#### 5.1.1 Onboarding and psychoeducation

In onboarding (session 1), users are oriented to the chatbot format and provided a general overview and rationale for the intervention. In the second session (psychoeducation), users learn about emotions and common reactions to stressful experiences through an exemplar fictional persona. Following this, they are encouraged to set goals within specific categories relating to their self-development including learning, relationships, and mood management. Based upon formative work with Jordanian youth, these domain categories were found to be relevant and acceptable to this age group of 18-21-year-old people. If a given specific goal does not relate, users are able to choose ‘something else’ and enter free text.

#### 5.1.2 Emotion Regulation

The user learns stress management strategies in sessions three and four. Specifically, in Session 3 slow breathing is taught as a simple tool to help reduce stress symptoms and enhance relaxation. In Session 4, the user learns about common problems when practicing slow breathing (e.g., feeling dizzy) and is offered ways to overcome these. Additionally, the user is given the opportunity to learn another stress management strategy, namely grounding using one’s five senses. Across both sessions, the user is encouraged to make a specific plan for practise by choosing a time of day for use of the techniques (e.g., mornings, lunchtimes, afternoons or evenings).

#### 5.1.3 Behavioural Activation

By choosing to follow one of two possible character stories, the user learns about the impact of actions on mood. These characters relay the impact of their mood on their activity levels, and ways they have overcome stressful periods with specific action taken in their lives. Across Sessions 5 and 6, the user is encouraged to choose a specific activity they can try out during times of stress that involve an experience of mastery, pleasure and/or social connection. Following this, the user learns to break down activity planning whereby they are guided to consider the ‘smallest first steps’ one would need to take when carrying out a chosen activity. Across these sessions, the user is encouraged by the bot to set a time of day during which time they will implement their chosen activity.

#### 5.1.4 Managing Problems

In Session 7, the user learns about a simple ‘Stop-Think-Go’ strategy to manage practical problems. Once again, the user chooses between one of two character stories to learn how to implement this strategy for common problems such as financial stress, unemployment, and relationship conflict. In the first step, the user guides the chosen character to pick one small problem, break down this problem into several parts, and in turn choose one small part (of a bigger problem) to which they can apply this strategy. In the second step, the character demonstrates by seeking support from a trusted other, they were able to generate possible solutions as well as brainstorm solutions of their own. Finally in the third step, the user encourages the character to carry out a specific solution and further learns about the impact of this action on the management of the problem (e.g., feelings of relief, agency). The bot then encourages the user to apply the ‘Stop-Think-Go’ strategy to one of their own problems and is guided by the bot on this.

#### 5.1.5 Supportive Self-talk

Across Sessions 8 and 9, the concept of self-talk during experiences of stress is introduced and the user is supported to think of alternative ways to view stressful situations. This is done by the user choosing to follow one of two possible character stories.

Specifically, the chosen characters talk about key biases that can arise in their thinking during times of stress (e.g., a friend not responding to their text messages), and how this can skew their reading of stressful experiences in a manner that worsens their stress and mood (e.g., ‘my friend doesn’t like me’). The character uses an analogy of “glasses” and guides the user to identify specific ‘disaster and negative glasses’ that one can wear during periods of stress, and how to refocus towards more balanced self-talk through asking themselves helpful questions (e.g., Will it matter in a year’s time?) that in turn help ‘remove’ these glasses.

Finally, the user is encouraged to notice their own self talk and related “glasses” that they may be wearing, and in turn practise helpful self-talk questions.

#### 5.1.6 Relapse Prevention

In Session 10, the bot congratulates the user on reaching the end, and asks the user to reflect on what they found helpful and useful throughout the core sessions. Next, the bot guides the user to identify potential triggers for situations of stress in the future, and in turn consider an action plan for a future stressful situation as it relates to use of the strategies the user has learned in the program.

### 5.2 Consolidating user learning

Based on formative work in Jordan, additional content was developed to complement the core sessions and further consolidate user learning. Specifically, users are given access to a ‘toolbox’ feature within the chatbot that provides them with access to audio and videos from core sessions. Second, users can engage with a practice session (‘*Helping session*’) involving having a conversation with a fictional persona facing a specific emotional problem (i.e., suffering from negative self-talk). In this session, users are guided to identify the presenting issue, and in turn implement learned techniques (e.g., supportive self-talk, manage problems) by taking an active role in offering suggestions to help the fictional persona to manage their presenting issue (e.g., what is the smallest first step to take). The session ends with users deriving their own action plan in the event they are faced with a similar presenting issue.

### 5.3 Access and Privacy

The chatbot was designed for access by smartphone or similar device where both a phone number and email could be used to access the platform. This meant that young people without a personal phone but with access to a device elsewhere (e.g., through a community centre) could access the intervention. Access to the chatbot requires a login and password created by the user. Data gathered is stored on a secure server. An internet connection is required to use the chatbot, whereby users can continue a conversation where they left in the event of technical difficulties (e.g., unstable internet connection).

## 6 Discussion

This paper describes the systematic development of an evidence-based chatbot designed to support distressed adolescents and young adults across LMIC settings using extensive fieldwork. The intervention was shaped by iterative testing rounds with end-users, community members, psychological intervention experts, and researchers. The design process utilised a HCD framework and led to the production of an automated chatbot (without natural language processing features) teaching core transdiagnostic CBT strategies in an interactive and engaging manner. The STARS chatbot was tested in a pilot randomised controlled trial in Jordan with outcomes relating to feasibility and acceptability (de Graaff et al., submitted). A fully powered test of the STARS chatbot is currently underway in Jordan (Akhtar et al., 2024). The strength of the STARS chatbot is in its co-development throughout all stages with key stakeholders within a human centred design framework. Addressing stigma was a key focus in the design of the chatbot where using language and examples that were non-diagnostic and relatable helped to tackle barriers to engagement among adolescents and young adults. This HCD approach to the development of STARS arguably increases its acceptability and scalability as end-users experience the intervention as it fits into their context, needs and conceptualisations of mental health. This is contrasted to currently available psychological interventions for youth including children and adolescents in LMIC settings that were developed within a primarily ‘top-down’ framework (Bryant et al., 2022; Hamdani et al., 2024). Whilst there is tremendous potential for digital chatbot interventions to circumvent typical barriers to youth seeking help (i.e., stigma, limited access to specialists), there are further considerations relating to guidance that must be considered. Across phases 1 and 2, end-users reported some barriers to engagement relating to privacy, confidentiality, and the need for support from trusted and learned (or ‘wise’) individuals. Whilst some of this was addressed in the automated chatbot; pairing a chatbot with human guidance may further address some of these issues (Schäfer et al., 2024). The proposed guidance model for use with the chatbot is based on a forthcoming WHO manual outlining a guided self-help approach which has been used with digital mental health interventions implemented in both middle-income and high-income countries (Cuijpers et al., 2022; Mediavilla et al., 2023).

Within the proposed framework, end-users are offered five, weekly 15-minute telephone-calls provided by a trained and supervised non-specialist helper complemented with messaging contact (e.g., what’s app or email for the purpose of scheduling). The structure of each weekly call includes (i) conducting a brief review of the week; (ii) reviewing sessions accessed by the user and their practise of learned techniques; and (iii) ways to encourage and support continued use of the chatbot. Importantly, the content of the helper training is designed to be independent of the chatbot content, such that guidance can be provided with varying pacing as suited to the needs of the user, whilst still adhering to the five-contact protocol. Helpers are also trained in basic helping skills (e.g., active listening, empathy) to respond to other problems that may be raised by users (e.g., financial support) and ways to ensure safety (e.g., identifying imminent risk of suicide) during use of the STARS chatbot.

To this end, helpers need to be trained to guide the user to identify strategies learned in STARS that could be applied to their presenting issue (e.g., coping with a physical health problem), and can accordingly be provided with other resources as needed (e.g., primary health care referrals). Helpers receive ongoing case supervision by a qualified mental health staff member (e.g., clinical psychologist).

### 6.1 Potential for adaptation and future directions

STARS was initially designed for use with existing non-AI chatbot systems that use technologies including applications, websites, and messaging platforms. These platforms typically require low amounts of data with the only requirement being an internet connection. For any future implementation, the content of conversational scripts will need to undergo adaptation including user and expert feedback to ensure the fictional personas, language, multimedia content, GIFs and emojis are culturally suited for the target user group. Further tests of this intervention may involve testing it as unguided or with more limited guidance to suit differently resourced contexts. An example variation may involve tests of this chatbot intervention with the current proposed five weekly support calls against a more limited form (i.e., ad hoc guidance). Such adaptations will provide important insight into the level of guidance needed and accepted by young people in each setting.

Additionally, recent rapid advancements of artificial intelligence (AI) have uncovered a space for using machine learning methods in digital mental health care including AI-assisted chatbots (Zhong et al., 2024). Future adaptations of the STARS chatbot may be enhanced by leveraging some degree of natural language processing (NLP) in the method by which content is delivered to users. For example, user experience with pre-set quick reply options throughout phases 1 and 2 indicated a preference for more personalised interactions with the chatbot, and this is one aspect of the chatbot that could potentially be refined with NLP features. A recent systematic review and meta-analysis of AI-based conversational agents in mental health care indicate that generative AI-based interventions can significantly reduce psychological distress amongst clinical populations relative to rule-based chatbots (Li et al., 2023). Considerations for such adaptations however must be carefully designed and balanced with the fact that generative AI methods carry some risks insofar as their unpredictable nature in leading to potentially negative outcomes (Luxton, 2020).

## 7 Conclusion

The STARS intervention has been designed with a focus that the content and scripts can be made publicly available by WHO following tests of its efficacy. Once efficacy is confirmed, the intervention can be made available – after adaptation - to wide ranging age groups including adolescents to young adults. As a novel non-AI based chatbot intervention, this work holds promise as a first step for youth seeking help within the context of structural and psychological barriers often surrounding them in LMICs. In particular, the anonymity that is enabled in the use of this intervention is ideally placed for this demographic who often report difficulties with stigma.

## Ethics

The current study received ethical approval from the School of Nursing, University of Jordan (PF.22.9 on March 23, 2022).

## Data Availability

N/A

## Acknowledgements

The authors wish to thank Dr Mark van Ommeren for supporting the development of this project.

